# Prevalence and Transmission of SARS-CoV-2 in Childcare Facilities: A Longitudinal Study

**DOI:** 10.1101/2021.04.16.21255616

**Authors:** Luise Haag, Judith Blankenburg, Manja Unrath, Johanna Grabietz, Elisabeth Kahre, Lukas Galow, Josephine Schneider, Alexander H. Dalpke, Christian Lück, Leo Büttner, Reinhard Berner, Jakob P. Armann

**Author notes:** **Corresponding Author:** Jakob P. Armann, Department of Pediatrics, University Hospital and Medical Faculty Carl Gustav Carus, Technische Universität Dresden, Fetscherstrasse 74, 01307 Dresden, Germany. Contributed equally as co-first authors (Luise Haag, Judith Blankenburg). **Conflict of Interest Disclosure:** Alexander H. Dalpke, Reinhard Berner and Jakob P. Armann report grants from the Federal State of Saxony during the conduct of the study. The other authors have no conflicts of interest to disclose. **Role of the Funding Source:** The funder of the study had no role in the study design, data collection, data analysis, data interpretation, or writing of the report. **Clinical Trial Registration:** KiTaCoviDD19: SARS-CoV-2 Surveillance im Vorschulalter (COVID-19), DRKS00022729, https://www.drks.de/drks_web/navigate.do?navigationId=trial.HTML&TRIAL_ID=DRKS00022729. **Data Sharing:** Deidentified individual participant data (including data dictionaries) will be made available, in addition to study protocols, the statistical analysis plan, and the informed consent form. The data will be made available upon publication to researchers who provide a methodologically sound proposal for use in achieving the goals of the approved proposal. Proposals should be submitted to. **Contributors’ Statement:** Dr Armann conceptualized and designed the study, wrote the protocol, collected samples, analyzed and verified the data, drafted the initial manuscript, and reviewed and revised the manuscript. Ms Haag and Ms Blankenburg collected samples, analyzed and verified the data, drafted the initial manuscript, and reviewed and revised the manuscript. Prof Berner conceptualized and designed the study, wrote the protocol, and reviewed and revised the manuscript. Prof Dalpke conceptualized and designed the study, wrote the protocol, performed laboratory analyses, and reviewed and revised the manuscript. Drs Lück and Büttner performed laboratory analyses, and reviewed and revised the manuscript. Dr Galow and Ms Unrath conceptualized and designed the study, wrote the protocol, collected samples, and reviewed and revised the manuscript. Dr Kahre, Ms Schneider and Ms Grabietz collected samples, and reviewed and revised the manuscript. All authors approved the final manuscript as submitted and agree to be accountable for all aspects of the work.

## Abstract

**Objectives:** Previous data indicate that children might play a less crucial role in SARS-CoV-2 transmission than initially assumed. We conducted a study to gain further knowledge on prevalence, transmission and spread of SARS-CoV-2 among preschool children, their parents and caretakers.

**Methods:** Children, their parents and care givers in 14 childcare facilities in Dresden, Saxony/ Germany were invited to participate in the KiTaCoviDD19-study between July 2020 and January 2021. Seroprevalence of SARS-CoV-2 antibodies was assessed up to 4 times during the study period in all participating adults and personal characteristics as well as epidemiological information of personal SARS-CoV-2 history were obtained. Stool viral shedding of SARS-CoV-2 was analyzed every 2-4 weeks in all participating children.

**Results:** In total, 318 children, 299 parents and 233 childcare workers were enrolled. The percentage of seropositive adults and SARS-CoV-2 positive detected children rose considerably by January 2021. However, the rate of SARS-CoV-2 positive children was considerably lower than the rate of seropositive adults. Overall, we detected a maximum of three connected cases in children. About 50% of SARS-CoV-2 infections in children could not be connected to a secondary case within our study population.

**Conclusion:** The study could not provide evidence for a relevant asymptomatic (“silent”) spread of SARS-CoV-2 in childcare facilities, neither in a low nor a high prevalence setting. This finding adds to the evidence that childcare and educational settings do not play a crucial role in driving the SARS-CoV-2 pandemic.

**Table of Contents Summary:** This longitudinal study among children, parents and childcare workers provides further insight on SARS-CoV-2 prevalence and transmission within childcare facilities.

**What’s Known on This Subject:** Based on age distribution of SARS-CoV-2 infections and previous data of very limited spread of COVID-19 among primary and secondary schools there is reason to believe that children play a less crucial role in SARS-CoV-2 transmission than initially assumed.

**What This Study Adds:** Previously published studies focus mainly on SARS-CoV-2 transmission in schools. This longitudinal study provides information on prevalence, transmission and spread of SARS-CoV-2 within childcare facilities during low- and high-prevalence settings.

## Introduction

Since the Beginning of COVID-19 pandemic ^1^ school and childcare closures are one of the main strategies to limit transmission. These measures are based on the assumption that children play a similar role in transmitting SARS-CoV-2 as they do in transmitting influenza and that schools and childcare facilities closures effectively lower the overall transmission rate ^2^.

However, most countries, including Germany, report a much lower proportion of cases in children compared to their population size ^3–5^. In addition, COVID-19 mostly leads to no or mild symptoms in children and presents a low risk of a serious course of disease to children, whereas the impact of school closures and limited social interactions on children’s mental health is becoming increasingly apparent ^6,7^.

While several studies in the UK and Norway could only identify a very limited spread of COVID-19 in primary and secondary schools ^8,9^ similar studies in preschools and childcare facilitis are lacking so far. Given the difficulty of implementing relevant distancing and hygiene meassures in this age group SARS-CoV-2 transmission in childcare facilities are of particular interest.

This study ivestigates SARS-CoV-2 transmission in childcare facilities attended by children between 1 and 6 years of age.

Mitigation strategies implemented by the Federal State of Saxony included a mask mandate for parents and limited access during drop-off and pick-up times. There was no recommendation for childcare workers or children to wear masks. Starting on December 14^th^, childcare facilities limited their service to emergency care for children of essential workers. Individuals with a confirmed SARS-CoV infection, close contact to an infected individual or individuals with respiratory symptoms with or without fever were not allowed to attend daycare facilities during the study period

## Methods

### Study Design

Children, their parents and childcare workers in 14 childcare facilities in Dresden, Saxony/ Germany were invited to participate in the KiTaCoviDD19-study.

After informed consent, 5 mL of peripheral venous blood were collected from adult participants and serological testing for IgG-antibodies against SARS-CoV-2 was conducted. Parents were asked to collect stool samples from their children every two weeks. Excretion of SARS-CoV2, as mostly non-infectious particle, is known to occur in infected people, especially children, for at least two weeks ^10^. This was used as an easy-to-obtain specimen in the childcare population. These stool samples were tested by PCR to detect stool excretion of SARS-CoV-2 RNA. All blood and stool samples were collected between July 2020 and January 2021.

In addition, data on age, household size, comorbidities, regular medication and previously diagnosed SARS-CoV-2 infections in participants or their household contacts quarantine episodes, utilization of daycare during lockdown, number of contacts other than household-contacts and the occurrence of respiratory symptoms were collected.

### Laboratory Analysis

Stool samples were frozen at −80°C and were thawed earliest after 4 weeks of storage. According to study protocol, initially, every second stool sample was examined. If at least one stool sample within one childcare facility tested positive, all samples from this institution collected two weeks earlier and later were tested as well.

The liquid handling system MicroLab Nimbus (Seegene, Duesseldorf, Germany) was used for nucleic acid extraction using a modified protocol supplied by the manufacturer (spatula point of stool in 1ml of ASL buffer, Qiagen/ mixed thoroughly and incubated for at least 10min/ centrifuged at 16.060g for 2 min/ supernatant was used for extraction procedure) as well as for PCR setup. Extraction was followed by real time reverse transcriptase polymerase chain reaction (RT-PCR) using Seegene’s Allplex™ 2019-nCoV assay. This assay detects three target regions within the genome of SARS-CoV-2, namely the E gene, RdRP gene and N gene. The automatic calculation software supplied by Seegene was used for interpretation of the results. A specimen was considered positive in case of at least one detected target region. In each test series a stool specimen from a patient with confirmed SARS-CoV-2 infection was run in parallel as a positive control for correct extraction and PCR procedure. In case of test inhibition (internal control) the assay was repeated once.

Blood samples were assessed for SARS-CoV-2 IgG antibodies using a commercially available chemiluminescence immunoassay (CLIA) technology for the quantitative determination of anti-S1 and anti-S2 specific IgG antibodies to SARS-CoV-2 (Diasorin LIAISON® SARS-CoV-2 S1/S2 IgG Assay). Antibody levels > 15.0 AU/ml were considered positive and levels between 12.0 and 15.0 AU/ml were considered equivocal.

All samples with a positive or equivocal LIAISON® test result, as well as all samples from participants with a PCR-confirmed SARS-CoV-2 infection, were tested with two additional serological tests (chemiluminescent microparticle immunoassay (CMIA) intended for the qualitative detection of IgG antibodies to the nucleocapsid protein of SARS-CoV-2 (Abbott Diagnostics® ARCHITECT SARS-CoV-2 IgG) and an ELISA detecting IgG against the S1 domain of the SARS-CoV-2 spike protein (Euroimmun® Anti-SARS-CoV-2 ELISA)).

### Definitions

Participants with detectable antibodies in at least two assays were considered seropositive. The study period was divided into a low-prevalence phase (July 15^th^, 2020 – November 15^th^, 2020) in which cumulative reported cases tripled in Dresden and a high-prevalence phase (November 16^th^, 2020 – January 31^st^, 2021) in which cumulative cases increased more than 10-fold (*figure 1*). Only children with at least two donated stool samples in the respectively defined periods were included in the analysis. 232 out of 318 children (73.0%) met these criteria in the low prevalence phase and 222 out of 318 children (69.8%) in the high prevalence phase (*table 1*). No positive stool sample was excluded by these criteria.

**Figure 1:**
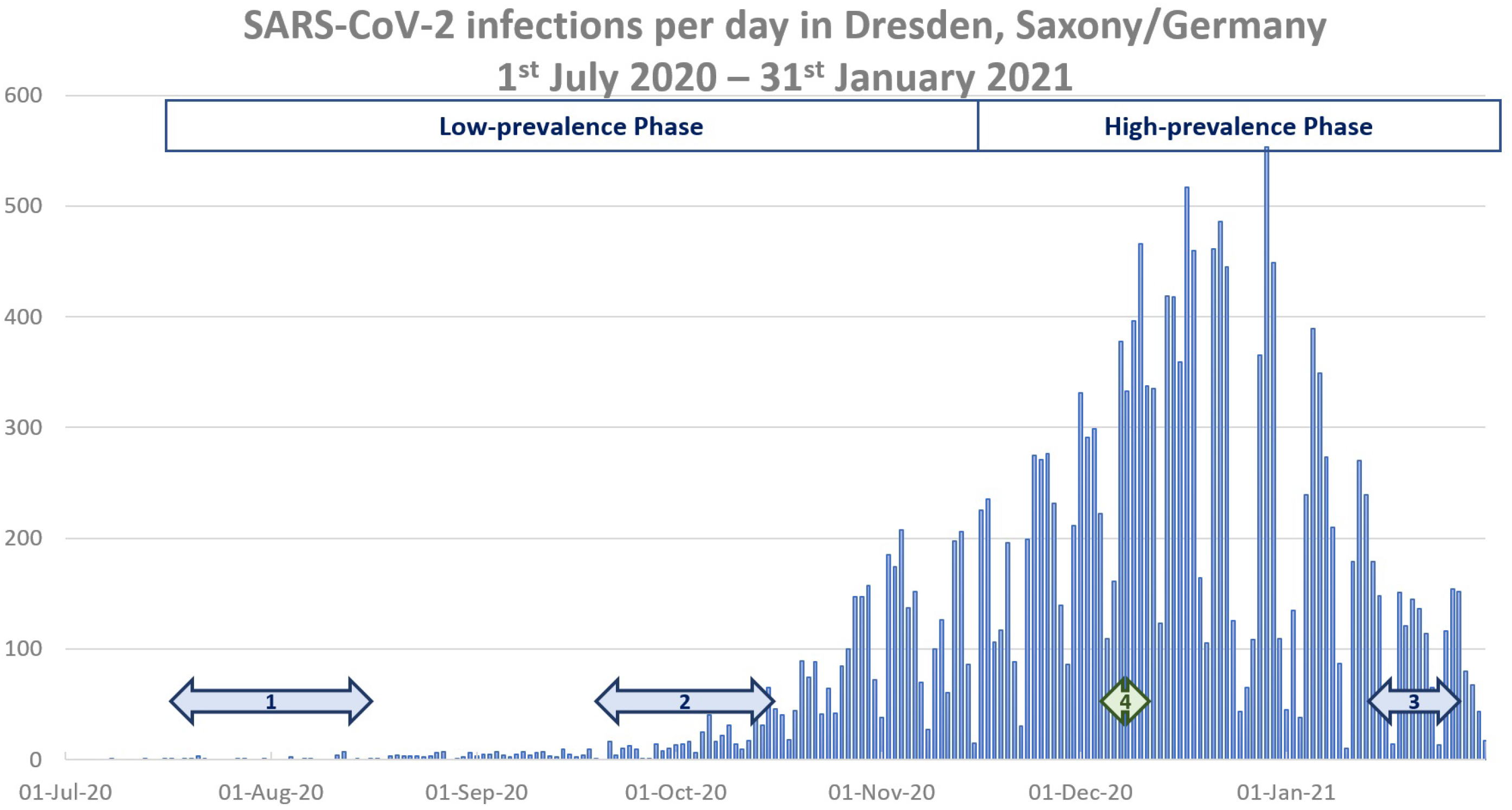
Timeline of serological testing (1: baseline, 2: 2^nd^ serological testing, 3: 3^rd^ serological testing, 4: additional serological testing in December 2020) and reported numbers of SARS-CoV-2 infections in Dresden, Saxony/Germany^15^

**Table 1:** Participants during low- and high-prevalence phase

|  | **Baseline** | **Low-prevalence Phase** | **High-prevalence Phase** |
| --- | --- | --- | --- |
| **Parents (n)** |  | 243 | 236 |
| **Childcare workers (n)** |  | 200 | 187 |
| **Children (n)** |  | 232 | 222 |

Cases within one facility were considered as possibly linked if there was an epidemiological and temporal association. Epidemiological association was defined as at least two SARS-CoV-2 infections within the same institution. Temporal association was defined as cases occurring within the infectious period of each other. This period is defined as 48 hours before onset of symptoms or positive PCR-test until 14 days after onset of symptoms. For stool samples we extended this period to 14 days before and after the positive sample was collected. In cases of consecutive positive samples, only the date of the first positive stool sample was taken into consideration as previous studies have shown that SARS-CoV-2 RNA can be shed through stool for multiple weeks whereas the infectious period is usually limited to the first 10 days ^11^.

### Statistical Analysis

Analyses were performed using IBM SPSS 25.0 and Microsoft Excel 2010. Fisher’s exact test was used to determine categorical variables for the statistical analysis. P-values ≤ 0.05 were defined as statistically significant.

### Approval

The KitaCoviDD19-study which was approved by the Ethics Committee of the Technische Universität (TU) Dresden (BO-EK-180052020), registered on July 15^th^, 2020 and assigned the clinical trial number DRKS00022729.

## Results

### Study population/ Demographics

318 children, 299 parents and 233 childcare workers in 14 childcare facilities were enrolled - representing 72% (50-100%) of all staff members and 18% (12-27%) of children attending these institutions (*table 1*). For 314 out of 318 children (99%) corresponding parental serostatus was available. 38 (12%) were siblings.

The median age of the children was 4 (IQR 2-5), their parents 37 (IQR 34-40) and childcare workers 39 (IQR 32-49). The median household size for children and parents was 4 (IQR 3-4) and 3 (IQR 2-3) for the participating childcare workers. (*table 2*.)

**Table 2:** Baseline Demographic Data; IQR: Interquartile Range

|  | **Childcare workers** | **Parents** | **Children** |
| --- | --- | --- | --- |
| **Number of participants** | 233 | 299 | 318 |
| **Age, median (IQR)** | 39 (32-49) | 37 (32-40) | 4 (2-5) |
| **Sex - female, %** | 88.2 | 68.2 | 49.5 |
| **Household size, median (IQR)** | 3 (2-3) | 4 (3-4) |  |

### Low-prevalence period

At baseline, none of the participants were SARS-CoV-2 seropositive. In the low-prevalence phase until mid-November 2 study participants, one childcare worker (1/154 – 0.7%) and one parent (1/196 – 0.5%) became seropositive. Both participants reported a known PCR-confirmed infection. They did not attend the same childcare facility.

During the same period, we detected 2 positive stool samples, resulting in a cumulative prevalence of 2 out of 232 children (0.9%) SARS-CoV-2 positive children (*table 3*). These children attended different childcare facilities. One case was not previously detected, however there was no epidemiological linked case in the same institution. The other child was symptomatic and SARS-CoV-2 infection was detected by PCR at that time. One epidemiological linked case occurred in the same institution. In both cases, the parents did not have detectable antibodies, nor did they report positive PCR-testing for SARS-CoV-2.

**Table 3:** Seropositive adult study-participants and SARS-CoV-2 positive children during low- and high-prevalence phase

|  | **Low-prevalence Phase** | **High-prevalence Phase** |
| --- | --- | --- |
| **All seropositive participants** | 2/443 (0.5%) | 48/424 (11.3%) |
| **Seropositive parents** | 1/243 (0.4%) | 25/236 (10.6%) |
| **Seropositive childcare workers** | 1/200 (0.5%) | 23/187 (12.3%) |
| **SARS-CoV-2 positive children** | 2/232 (0.9%) | 15/222 (6.8%) |

### High-prevalence period

At the end of January 2021 - after the second wave of the pandemic - 25 out of 236 (10.6%) parents and 23 out of 187 (12.3%) childcare workers were seropositive (*table 3*). The difference between the two groups showed no statistical significance (p=0.64).

Seroprevalence in parents whose children remained in emergency care during the strict lockdown was significantly higher (12/64 (18.8%)) than parents whose children did not visit the facility during this time (13/152 (8.6%); p=0.038). Seroprevalence in childcare workers assigned mainly administrative was significantly higher (10/48 (20.8%)) than those with mainly childcare related duties (8/99 (8.1%) p=0.034

Eleven seropositive participants (2 childcare workers and 9 parents) had an additional serum sample taken mid-December. Three out of 11 (27.3%) were already seropositive at this time while 8 out of 11 (72.7%) seroconverted during the strict lockdown starting December 14^th^, 2020.

A total of 68 out of 187 childcare workers (36.4%) and 93 out of 236 parents (39.4%) reported symptoms of an upper respiratory tract infection during the study period.

### Seropositive parents and childcare workers

20 out of 25 seropositive parents (80.0%) had been previously tested positive for SARS-CoV-2 and one reported a confirmed SARS-CoV-2 positive household member. Similarly, 18 out of 23 childcare workers (78.3%) had a personal SARS-CoV-2 history, and one childcare worker had a respective household history. The ratio of undetected to detected SARS-CoV-2 infections did not differ significantly between parents (4/21 (0.19)) and childcare workers (4/19 (0.21)).

Ten out of 25 seropositive parents (40.0%) and 5 out of 23 seropositive childcare workers (21.7%) named a confirmed SARS-CoV-2 contact outside the childcare facility as source of infection.

### SARS-CoV-2 positive children

During the second study period 15 out of 222 children (6.8%) had at least one SARS-CoV-2 positive stool sample. These cases occurred in 8 different institutions. In 4 childcare facilities a possible epidemiological link between a maximum of 3 children each could be identified. In the remaining 6 of 14 cases there was no connection to other children with SARS-CoV-2 positive stool samples.

Altogether, SARS-CoV-2 RNA was detected in a total of 22 out of 1168 analysed stool samples, belonging to 17 different children. 5 out of 17 children had two consecutive positive samples. In 8 out of 17 (47%) cases a personal or household contact PCR-confirmed SARS-CoV-2 infection was reported, leading to a ratio of 1.125 (9:8) of undetected to detected SARS-CoV-2 infections in the participating children.

64 out of 219 parents (29.2%) reported that their children visited the childcare facility during the strict lockdown starting on December 14^th^, 2020. Cumulative SARS-CoV-2 prevalence was not significantly between children attending emergency care and those who did not (4/64 (6.3%) vs 13/152 (8.6%); p=0.78).

### SARS-CoV-2 transmission in childcare facilities

The 17 SARS-CoV-2 positive children attended 8 different childcare facilities, in the remaining 6 facilities no positive stool samples were detected. The maximum number of children with a SARS-CoV-2 positive stool sample per center was 4. 2 out of 17 children with stool viral shedding had siblings who were enrolled in this study as well. In both cases the stool samples of the respective sibling yielded negative results. 5 out of 17 children (29.4%) did not visit the childcare center prior to or during their viral shedding, thereby eliminating the possibility of an infection connected to their childcare center.

Six out of 17 parents (35.3%) with a SARS-CoV-2 infected child were seropositive, 3 parents did not undergo serological testing after their children’s positive stool sample and 8 out of 17 (47.1%) were seronegative.

12 of the 17 children with a SARS-CoV-2-positive stool sample attended their childcare facility at the time of viral shedding. This affected 5 institutions. In 7 cases at 4 institutions at least one epidemiologically associated SARS-CoV-2 case could be detected. In 5 out of 12 cases there was no epidemiological associated SARS-CoV-2 case within the same facility.

Regarding the occurrence of epidemiological linked transmissions within the participating institutions we detected at least one SARS-CoV-2 positive participant in 13 out of 14 (92.8%) childcare facilities. In 8 out of 14 childcare facilities (57.1%) none or only isolated cases without possible epidemiological association occurred. In 3 out of 14 (21.4%) there was a maximum of 2 SARS-CoV-2 infections with a possible epidemiological link while in 3 out of 14 facilities (21.4%) there were outbreaks involving at least 3 (3-10) cases. In all outbreaks positive stool samples in children could be detected *(figure 2)*.

**Figure 2:**
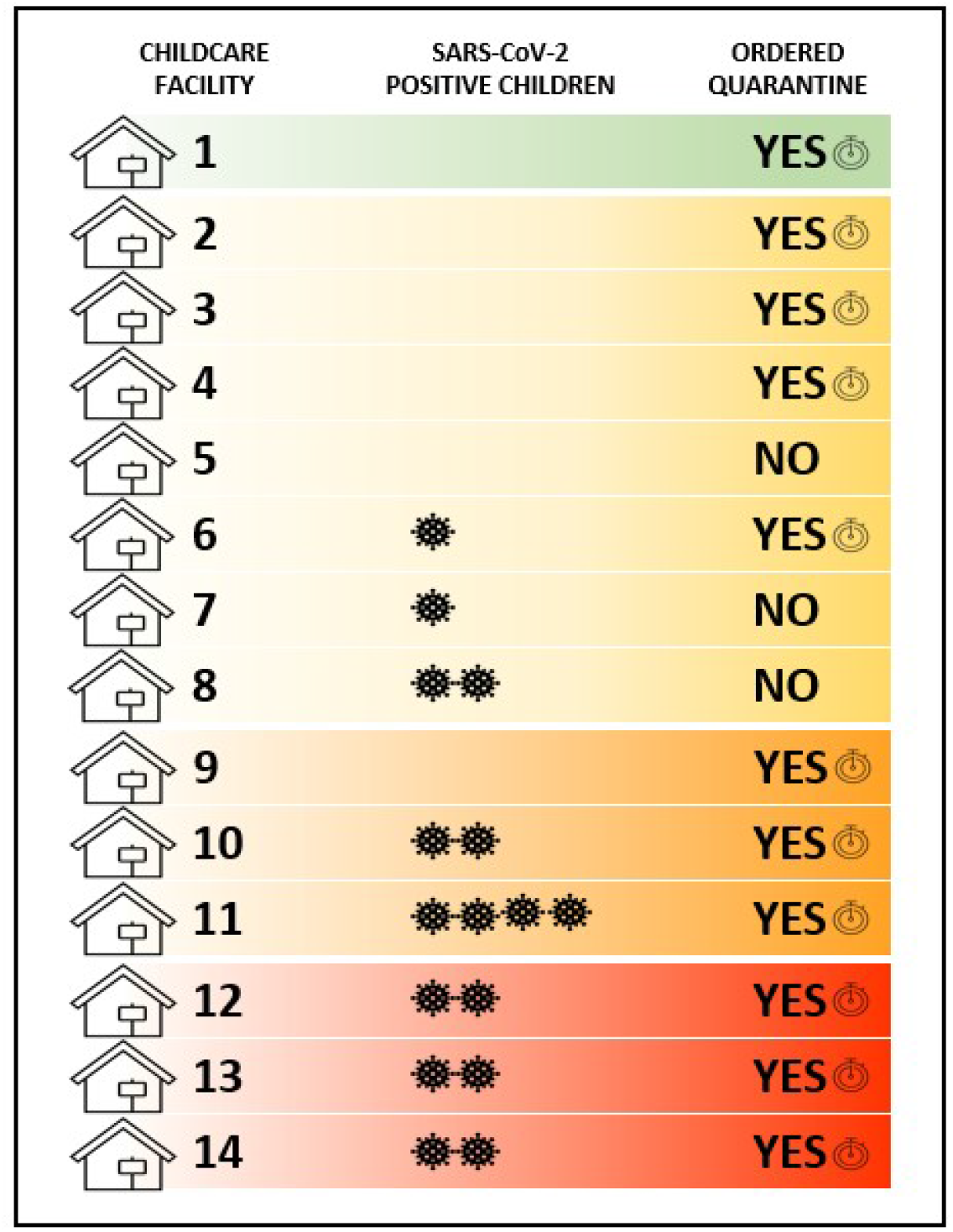
Positive SARS-CoV-2 infected children and the occurrence of epidemiological linked transmissions within the single participating childcare facilities (*green* – no epidemiological link detected, *yellow* – single cases without epidemiological link detected, *orange* – 2 epidemiologically linked cases detected, *red* – outbreaks with > 2 epidemiologically linked cases detected, *stopwatch* – officially mandated quarantine by local health department during study period)

7/9 children with undetected SARS-CoV-2 infection attended their childcare facilities during viral shedding. In 3 cases we could detect none or maximal one possibly linked case. 4 cases were linked to outbreaks in two different childcare facilities. Given the retrospective study design, we cannot make assumptions about the index case of the outbreaks though.

### Quarantine

Eleven of the 14 participating childcare facilities had at least one episode of mandated quarantine by the local health department due to a PCR-confirmed SARS-CoV-2 infection at their institution. In one of the childcare facilities with an ordered quarantine episode we could not detect any positive stool sample or seropositive adults. At all 3 institutions with detected outbreaks quarantine was ordered accordingly. In 2 of the remaining 3 childcare facilities without quarantine episodes we detected a total of 3 SARS-CoV2-positive stool samples with part of the children attending their childcare facility during time of stool viral shedding. In all three childcare facilities without quarantine episodes, we could only detect isolated cases of SARS-CoV-2 with no epidemiological link.

## Discussion

The rate of seropositive participants during the low-prevalence phase following the first wave in the Federal State of Saxony/Germany was very low, with only 2 detected cases. Both of these participants previously knew of their infections, quarantine warranties were issued accordingly, and we could not find connected cases to either participant. This supports the assumption that the current testing and quarantine methods in Germany during a low-prevalence phase were effective and could successfully prevent an undetected spread of SARS-CoV-2. In accordance, we could only find evidence for one undetected infection in a child during this period. Even though this child visited the childcare facility and no special hygiene or distancing measures were taken neither by the parents nor by the childcare workers, we did not detect a secondary case.

The increasing SARS-CoV-2 cases during the second wave in the general population was mirrored in our study population. While the rate of seropositive participants rose considerably by January 2021, this increase was proportional to the increase within the general population in Dresden, Germany. The number of undetected infections detected only by antibody testing continued to be lower than previously assumed by some authors. (DOI: 10.1126/science.abb3221).We could not detect a significant difference between the rate of seropositive parents and childcare workers. The percentage of SARS-CoV-2 positive children within the high-prevalence study phase was considerably lower than the rate of seropositive adults. This finding is supported by previous studies that also reported lower infection rates in children ^5,12^.

Over the whole study period, we detected a maximum of three connected cases in children, further strengthening the hypothesis that childcare facilities are not a major source of uncontrolled clusters ^13^. Noteworthy as well was the fact that in two cases an infection among siblings was detected, in both cases the other sibling did not test positive in the corresponding stool samples.

About 50% of SARS-CoV-2 infections in children could not be connected to a secondary case within our study population.

While the rate of participating children only amounted to approximately 20%, we included an average of 72% of childcare workers. Due to this high participation rate, we are confident to have a reliable amount of samples analysed to evaluate the rate of SARS-CoV-2 infections among this group. We could not detect a significantly larger rate of infected childcare workers compared to the participating parents. While parents were significantly more often seropositive when their children visited the childcare facility during the lockdown period, this also implies a working environment without the possibility of home-office. This effect could therefore also be owed to the larger number of regular contacts within the work environment outside the household. This assumption is supported by the lower percentage of SARS-CoV-2 positive children in emergency care compared to those who stayed at home during the strict lockdown.

The percentage of seropositive childcare workers whose infection was possibly connected to their childcare facility was considerably higher than among seropositive parents. Considering the fact that there was a significantly higher rate of seropositive childcare workers with mainly administrative duties compared to those without, the higher infection rate in childcare workers compared to children as well as the extremely limited spread linked to undetected SARS-CoV-2 positive children suggests that transmission between adults within the childcare facilities occurs more frequently than between children and adults. Therefore, hygiene and distancing measures between childcare workers themselves might be the key measures in these institutions and more important than between children and workers.

There are several limitations to our study. We enrolled a limited number of participants in our study, thus it was possible that infections remained undetected. Moreover, a lower sensitivity of PCR-testing for SARS-CoV-2 in stool samples than in oro- and nasopharyngeal swaps might also contribute to underestimating the true numbers of infections ^14^.

However, the inclusion of low- as well as high-prevalence settings within the study period and the high participation rate of childcare workers in all facilities are strengths of our study. Additionally, the use of antibody testing instead of PCR testing reduces the possibility of missed undetected SARS-CoV-2 cases at least in parents and childcare workers.

## Conclusion

The study could not provide evidence for a relevant asymptomatic (“silent”) spread of SARS-CoV-2 in childcare facilities, despite the lack of hygiene or distancing measures in this age group. In addition, adults seem to transmit SARS-CoV-2 more frequently than children. These findings add to the evidence that childcare and educational settings do not play a crucial role in driving the SARS-CoV-2 pandemic.

## Data Availability

Deidentified individual participant data will be made available, in addition to study protocols, the statistical analysis plan, and the informed consent form. The data will be made available upon publication to researchers who provide a methodologically sound proposal for use in achieving the goals of the approved proposal. Proposals should be submitted to

## Abbreviations

COVID-19: Coronavirus disease 2019
ELISA: enzyme-linked immunosorbent assay
IgG: Immunoglobulin G
IQR: Interquartile Range
PCR: Polymerase Chain Reaction
SARS-CoV-2: severe acute respiratory syndrome coronavirus type 2

## Acknowledgements

We thank R. Fischer for excellent technical support.

